# The German Gestational Diabetes Study (PREG): Rationale, Methodology and Design

**DOI:** 10.1101/2021.10.10.21264499

**Authors:** Louise Fritsche, Julia Hummel, Robert Wagner, Dorina Löffler, Julia Hartkopf, Jürgen Machann, Johannes Hilberath, Konstantinos Kantartzis, Peter Jakubowski, Jan Pauluschke-Fröhlich, Sara Brucker, Sebastian Hörber, Hans-Ulrich Häring, Michael Roden, Annette Schürmann, Michele Solimena, Martin Hrabe de Angelis, Andreas Peter, Andreas L. Birkenfeld, Hubert Preissl, Andreas Fritsche, Martin Heni

## Abstract

**Introduction:** Since the introduction of the new International Association of the Diabetes and Pregnancy Study Groups (IADPSG) criteria for gestational diabetes mellitus (GDM) in 2012, diagnosis and treatment of GDM has improved. But even well-treated GDM might still have impact on long-term health of the mother and her offspring, though, this relation has not been conclusively studied yet.

**Methods:** The multicenter PREG study is designed to metabolically and phenotypically characterize women with a 75 g five-point oral glucose tolerance test (OGTT) during and repeatedly after pregnancy. The offspring of the study participants are followed up until adulthood with developmental tests and metabolic and epigenetic phenotyping in the PREG offspring study. By in-depth phenotyping of the mother and her offspring, we aim to elucidate the relationship of maternal hyperglycemia during pregnancy and adequate treatment and its impact on the long-term health for both.

**Ethics and dissemination:** The study protocol has been reviewed and approved by the ethics committee of the University Hospital Tübingen (protocol numbers 218/2012BO2 and 617/2020BO1), the ethics committee of the Technical University Dresden (protocol number EK263072013), the ethics committee of the medical school of the Heinrich Heine University Düsseldorf (protocol number 4051) and the ethics committee of the medical school of University of Leipzig (protocol number 038-15-09032015). The results will be disseminated through conference presentations and peer-reviewed publications.

**Registration:** The PREG study and the PREG offspring study are registered with Clinical Trials (ClinicalTrials.gov Identifiers: NCT04270578, NCT04722900).

**Strengths and limitations of this study:**

- The main strength of the multicenter PREG study are the in-depth phenotyping of mothers during pregnancy and repeatedly after delivery.
- Data acquisition and sample handling are done according to standard operating procedures in all study sites, thus, ensuring a high quality for each data point.
- A PREG biobank is set up and samples are available for researchers of the German Center for Diabetes Research (DZD).
- Children of the study participants are repeatedly examined to cover the period of childhood and adolescence.
- The PREG study is not planned as a population-based cohort but is enriched for GDM cases.

**Data Availability Statement:** All requests for data and materials will be promptly reviewed by the Data Access Steering Committee to verify whether the request is subject to any intellectual property or confidentiality obligations. Individual-level data may be subject to confidentiality. Any data and materials that can be shared will be released via a Material Transfer Agreement.

## Introduction

Gestational diabetes mellitus (GDM) is characterized by a defined degree of hyperglycemia that is first diagnosed during pregnancy. In 2008, the Hyperglycemia and Adverse Pregnancy Outcomes (HAPO) study [1] demonstrated the negative impact even of only moderately elevated blood glucose levels on pregnancy and birth outcome. Driven by these results, the International Association of the Diabetes and Pregnancy Study Group (IADPSG) recommended in 2010 new plasma glucose thresholds for the diagnosis of GDM with a 2 h 75 g oral glucose tolerance test (OGTT): fasting glucose 5.1 mmol/L, 1 h glucose 10 mmol/L and 2 h glucose 8.5 mmol/L [2]. In many countries, these GDM diagnostic criteria are in use and standardized screening procedures have been implemented. This approach enables the timely diagnosis of the majority of GDM cases. The prompt treatment has led to a drastic reduction of macrosomia and other unfavorable peripartal outcomes [3–6].

However, even if maternal glucose levels can be therapeutically controlled and, in most cases, normalize after delivery, GDM is not only a transient pregnancy-specific pathology. For the mother, GDM represents a window into her metabolic future [7]. Metabolic disturbances during pregnancy are predictive for women’s future risk of type 2 diabetes mellitus (T2DM) and cardiovascular diseases. This has been shown by a multitude of studies and meta analyses which demonstrated that GDM increases the risk to develop not only type 2 diabetes [8,9] but also the metabolic syndrome [10,11], cardiovascular disease [12] and some form of cancer [13]. A potential association with depression is currently controversially discussed [14,15].

For the offspring who had already been exposed to elevated glucose and nutrient availability by the time of GDM diagnosis, maternal GDM is associated with an elevated risk of adverse effects on (brain) development and metabolic health [16]. A number of studies show that GDM predisposes the offspring to become overweight and obese during childhood and adolescence [17]. Furthermore, disturbances of glucose metabolism in prepubertal children from GDM pregnancies have been reported [18]. Presumably via mechanisms termed fetal programing, high maternal glucose levels and glucose metabolism disorders cause epigenetic changes in the offspring that are detectable e.g. from placenta and blood [19,20]. The influence of GDM on the epigenome seems to be long-lasting as differential methylation patterns persist even in teenagers of mothers with increased glucose levels during pregnancy [21]. Adult offspring of mothers with GDM have a higher risk for glucose intolerance, insulin resistance [22] and T2DM [23]. Register studies furthermore suggest an adverse impact on cognitive development and intelligence [24,25]. Studies from our group demonstrated that the brain response of fetuses from mothers with GDM and insulin resistance is different from healthy controls, suggesting brain insulin resistance in these fetuses [26]. Furthermore, an association with attention deficit/hyperactivity disorder (ADHD) in children exposed to diabetes *in utero* has been described [27]. In addition to these potential central nervous effects, GDM appears to also affect the fetal autonomic nervous system [28].

Currently it is unclear whether standard GDM treatment is sufficient to fully prevent adverse long-term effects of GDM, such as obesity or disturbances of glucose metabolism in older children and adults. Some studies indicate that current GDM treatment approaches might not be sufficient to prevent increased childhood obesity [29,30] but might be beneficial for fasting glucose in female offspring age 5-10 years [31]. However, this has been investigated only by a limited number of studies and this issue needs further research in order to optimize GDM screening and therapeutic strategies during pregnancy.

### Transgenerational Diabetes

Genetic and environmental factors such as sedentary lifestyle and poor nutrition are discussed as major driver of the worldwide diabetes epidemic. Maternal hyperglycemia, i.e. GDM, might additionally drive the familial accumulation of T2DM via non-genetic inheritance mechanisms. Though, these mechanisms are currently incompletely understood. An oversupply of nutrients to the fetus may not only induce overgrowth via fetal hyperinsulinemia but may also introduce long-term changes in regulation of satiety, activity and endocrine system. Women with a family history of diabetes have a higher risk to develop GDM themselves, thus perpetuating this cycle of transgenerational diabetes.

### Aims

The detrimental effect of untreated GDM on the health of mother and child is undisputable, whereas the long-term effects of a well-treated GDM on mother and child are still largely unknown. Even if the immediate threat of fetal overgrowth and macrosomia could be dramatically reduced by nutritional intervention and medical treatment, fetal programing happens from start of pregnancy long before GDM is diagnosed. A treatment in the last trimester might not be sufficient to entirely counteract adverse maternal metabolic influences during the first two trimesters entirely.

By in-depth phenotyping of the mother during pregnancy and repeated follow up of her and her offspring, the PREG study provides an outstanding opportunity to elucidate the relationship of the maternal metabolism during pregnancy and its impact on the long-term health consequences for both.

## Methods

### Setting and study outline

The PREG study is a prospective multicenter cohort study conducted at four sites of the German Center for Diabetes Research (DZD) (Suppl. Table 1), coordinated by the Tübingen site. In all study sites, women are examined during pregnancy and up to four times in the decade after delivery. Birth outcome as well as offspring anthropometric data are collected. Cord blood and placenta are collected and the offspring cohort is investigated during subsequent visits (presently only at the Tübingen site).

The study outline is presented in Table 1. Women are examined during pregnancy as well as 1, 2, 5 and 10 years after delivery. Offspring are followed up at the age of 2, 6, 10, 14 and 17 years. Harmonized standard operating procedures are established for OGTT, spiroergometry, magnetic resonance imaging (MRI) and spectroscopy (MRS), sample generation, sample processing and sample storage across all study sites.

The PREG study and the PREG offspring study are registered with clinical trials (ClinicalTrials.gov Identifiers: NCT04270578, NCT04722900).

**Table 1.** Study outline

|  | Timepoint | OGTT | (Fasted) blood sample | Anthropometry | Continuous glucose measurment | Bioimpdance analysis | Liver ultrasound | Whole body MRI/ liver fat MRS | Spiroergometry | Developmental and intelligence tests | Questionnaires and food diary | Accelerometry |
| --- | --- | --- | --- | --- | --- | --- | --- | --- | --- | --- | --- | --- |
| During pregnancy | Gestational week 24-32 | X |  | X |  | X | X |  |  |  | X |  |
|  | Delivery |  | X<br>(Cord blood and maternal HbA1c) | X<br>(medical records) |  |  |  |  |  |  |  |  |
| Follow up mothers | 1 year | X |  | X |  | X |  | X | X |  | X |  |
|  | 2 years | X |  | X |  | X |  | X | X |  | X |  |
|  | 5 years | X |  | X |  | X |  | X | X |  | X |  |
|  | 10 years | X |  | X |  | X |  | X | X |  | X |  |
| Follow up children | 2 years |  |  | X |  | X |  |  |  | X | X |  |
|  | 6 years |  | X | X | X | X |  | X |  | X | X | X |
|  | 10 years |  | X | X | X | X |  | X |  | X | X | X |
|  | 14 years |  | X | X | X | X |  | X |  | X | X | X |
|  | 17 years |  | X | x | X | X |  | X |  | X | X | X |

### Recruitment

Eligible women are recruited via outpatient clinics for pregnant women, via diabetes outpatient clinic, via residential gynecologist practices and advertising leaflets / broadcast-emails. In Germany, GDM is currently mostly diagnosed with a two-step approach. All pregnant women between gestational week 24 and 28 are recommended to undergo a 50g glucose challenge test (GCT). This test can be performed at any time of day and does not require the women to be fasted. If the venous plasma glucose is ≥ 7.5 mmol/L at 1 h after 50 g glucose ingestion, a diagnostic 2h-75g-OGTT is performed on a separate day after an overnight fast. The 50g GCT is not part of the PREG study. Residential gynecologists refer patients with glucose concentrations above the threshold in the 50g GCT to the diabetes outpatient clinic where the women are offered participation in the PREG study. With this recruitment strategy, women with GDM are enriched in the PREG cohort and will therefore represent a high-risk group but will not be representative of the general population.

### Participants

Pregnant women are enrolled between gestational week 24+0 and 31+6. Additionally, women with a confirmed diagnosis of GDM in a previous pregnancy are eligible for the study, if detailed records of the previous diagnostic OGTT are available. Exclusion criteria are age < 18 years, preexisting type 1 or type 2 diabetes mellitus, estimated glomerular filtration rate (eGFR) < 60 ml/min/1.73m^2^ (MDRD-formula[32]), C-reactive protein (CRP) > 1mg/dL, transaminases > 2 times upper limit of the norm, preexisting cardiac conditions, weight loss >10% within 6 months prior to study enrollment, psychiatric disorders, chronic alcohol or drug abuse, blood glucose increasing or decreasing medication (e.g. steroids, oral antidiabetic agents, insulin). For subjects enrolling after delivery, additional exclusion criteria are current breastfeeding and pregnancy.

For each of the facultative measurements MRI/MRS and spiroergometry, there is a set of additional exclusion criteria each; for MRI/MRS: any ferromagnetic metals in or on the body, claustrophobia, impaired thermal sensor reception or increased sensitivity to heating, hearing loss or increased sensitivity to loud noises; for spiroergometry: cardiac morbidities (acute coronary syndrome, cardiac arrhythmias, congestive heart failure, acute carditis), history of pulmonary embolism, acute phlebothrombosis of lower extremities, hyperthyroidism or hypokalemia.

Exclusion criteria for offspring are severe malformations that preclude developmental tests.

### Ethics and dissemination

Informed written consent is obtained from every study participant and their legal guardian, respectively and the study is conducted in accordance to the declaration of Helsinki. The study protocol has been reviewed and approved by the ethics committee of the University Hospital Tübingen (protocol numbers 218/2012BO2 and 617/2020BO1), the ethics committee of the Technical University Dresden (protocol number EK263072013), the ethics committee of the medical school of the Heinrich Heine University Düsseldorf (protocol number 4051) and the ethics committee of the medical school of University of Leipzig (protocol number 038-15-09032015). The results will be disseminated through conference presentations and peer-reviewed publications.

A PREG biobank is set up and samples are available for researchers of the German Center for Diabetes Research (DZD) upon application and positive evaluation by the DZD Use and Access Committee.

### GDM diagnosis and treatment

GDM is diagnosed according to the national guidelines [33] of the German Diabetes Association (DDG) in accordance with the WHO and IADPSG [2] criteria. Patients with GDM are treated according to national guidelines [33] with nutritional counseling, self-monitoring of blood glucose, and insulin when necessary. This treatment is not part of the PREG study but is recorded in detail for each participant.

### OGTT and fasted blood sample

After a fasting period of at least 10 h, participants undergo a 5-point 2h oral glucose tolerance test (OGTT) with 75 g glucose during pregnancy as well as at all follow up visits (Table 1). Venous blood is taken from a venous catheter at 0, 30, 60, 90 and 120 minutes. Spot urine is collected before start of OGTT and immediately after the last blood drawing. In children (6-years and older), a blood sample is drawn in the morning after an overnight fast. Samples are immediately put on ice and are quickly processed or transferred to long term storage at -80°C, respectively.

### Laboratory measurements

From fasted blood samples, routine laboratory parameters, including HbA_1c,_ lipids, CRP, IL-6, cortisol, β-human chorionic gonadotropin, prolactin, estradiol, testosterone and sex hormone binding protein are measured (Suppl. Table 2). From each time point of OGTT glucose and non-esterified fatty acids are measured locally at each study site, while serum insulin and C-peptide are measured centrally at the Tübingen site by an immunoassay on ADVIA Centaur XP Immunoassay System (Siemens Healthcare Diagnostics, Erlangen, Germany). For batch analyses, plasma, serum and urine samples are stored at -80°C in the PREG-biobank (ethics board number for biobank: 141/2019BO1).

### DNA and RNA extraction

DNA is extracted from whole blood with appropriate steps of cell lysis, protein precipitation and washing. RNA is extracted from PAXGene (BD, Heidelberg, Germany). DNA and RNA are stored at - 80°C.

### Fetal magnetencephalography (fMEG) and magnetocardiography (fMCG)

In a subset of pregnant women, an additional measurement of fetal brain activity and heart rate is performed at the Tübingen site during the OGTT (ethics board number 339/2010BO1) [34]. Measurements are conducted with the special developed fetal MEG system SARA II (SQUID Array for Reproductive Assessment, VSM MedTech Ltd., Port Coquitlam, Canada). The device contains 156 integrated primary and 29 reference sensors (SQUID coils) to capture magnetic signals elicited by the fetal brain (fMEG) and heart (fMCG). The recording of biomagnetic signals is a unique tool to study the development of the human nervous system completely non-invasively *in utero* from as early as 28 weeks of gestation. Currently, only two devices of the SARA-type are installed worldwide (University of Arkansas for Medical Sciences, Little Rock, USA and fMEG Center of the University of Tübingen, Germany).

### Sonography of liver fat in pregnant women

The presence of liver fat accumulation is estimated using an ultrasonography of the liver by an experienced sonographer. The ultrasound examination is performed with a curved-array-transducer (34 Hz) and a CX50 POC ultrasound device (Philips, Hamburg, Germany). To quantify the degree of liver steatosis, a grading system is used to classify liver steatosis between grade I and III (Suppl. Table 3) [35]. In short, this classification is based on echogenicity, attenuation of sound and the visualization/differentiation of vessels. The echogenicity of the liver is normalized by comparing renal and hepatic parenchyma.

### Magnetic resonance imaging (MRI) and spectroscopy (MRS)

Non-pregnant adult participants undergo MR examinations on a 3 T whole body imager (Magnetom VIDA, Siemens Healthineers, Erlangen, Germany) for differentiation and quantification of whole-body adipose tissue (AT) compartments (by MRI) and determination of intrahepatic lipids (IHL) by MRS in the early morning after overnight fasting. As described in detail in [36], a T1-weighted fast spin echo technique – where AT appears distinctively brighter than lean tissue (LT) – is applied from feet to head with subjects lying in prone position with extended arms in a measuring time of about 20 minutes. Segmentation of AT and LT is performed by applying an automatic algorithm based on fuzzy-clustering and orthonormal snakes [37] within 2-3 minutes. Slight imperfections of segmentation due to inhomogeneous magnetic field distribution are manually corrected by visual inspection. From this, AT and LT of the lower extremities (feet to hips), abdominal AT (from hips to shoulders) and AT of the upper extremities (from shoulders to hands, including the head) are quantified. Abdominal AT is subdivided in visceral adipose tissue (VAT, ranging from hips to thoracic diaphragm) and subcutaneous AT (SCAT, including internal AT between thoracic diaphragm and shoulders). Localized proton MRS is performed in the posterior part of segment 7 in the liver applying a single voxel STEAM technique [36]. After morphologic MRI and careful shimming, 32 acquisitions are recorded from a volume of 3×3×2 cm^3^. (IHL content is given as the percentage of fat signals (including methylene and methyl) divided by the sum of water and fat signals.)

Children age 6 years and up are measured in the early morning after an overnight fast in a 3 T whole body imager (Magnetom VIDA, Siemens Healthineers, Erlangen, Germany). They are placed in supine position and covered with surface coils to ensure acceptable signal-to-noise ratio. First, an overview scan (fast-view, 15 seconds) is carried out, which is used for the exact positioning of the following images. Using a chemical-shift-selective recording technique (VIBE-Dixon), gapless fat- and water-selective images from the shoulders to the thighs are recorded in 2 blocks. The measurement time per block is 16 seconds, the measurements are made during an atomic stop in order to generate artifact-free images. After each acquisition, the patient table is moved to the position to be measured, since homogeneous images can only be generated in a narrow area around the isocenter of the magnetic field (approx. 50 cm). The fat distribution in the trunk of the body (visceral fat, subcutaneous fat) can be quantified from these tomograms. To quantify the intrahepatic lipid content (IHL), a spectrum from the posterior portion of segment 7 is acquired using volume-selective MR spectroscopy. The measurement takes approx. 1 minute and is carried out with continuous breathing with instructions. In addition, another imaging sequence (q-Dixon) comparable to that mentioned above is being applied, with the help of which ectopic lipid deposits in abdominal organs (liver, pancreas) can also be recorded. Measurement time is also 16 seconds during breathing stop. Measurement time in total, including preparation, amounts to approximately 15 minutes.

### Cord blood collection

From deliveries in the University Women’s Hospital Tübingen, cord blood is collected and quickly processed. Besides assessment of blood count and blood gas analysis, cord blood serum and EDTA-plasma samples are stored at -80°C for batch analyses.

### Anthropometry, body composition and blood pressure

Weight and height of participating women and their offspring are measured using a calibrated scale and stadiometer. From mothers, waist and hip are measured according to standard operating procedures with a tape measure. From children, height, weight, abdominal girth and head circumference are measured. Body composition is assessed by bioimpedance with an Akern BIA101 (SMT medical GmbH & Co, Wuerzburg, Germany) and calculated with Cyprus Version 2.7 for women (RJL Systems, Clinton Township, MI USA) and with BodyGramPRO software for children (SMT medical GmbH & Co, Würzburg). Blood pressure is measured with an automated sphygmomanometer with respective blood pressure cuff sizes for mothers and children (BOSO Carat Professional, Bosch + Sohn GmbH & Co, Jungingen, Germany).

### Spiroergometry

Nonpregnant women participate in a continuous, incremental exercise test to volitional exhaustion using a cycle ergometer (Ergometrics 800 S, Ergoline, Bitz, Germany). Oxygen consumption (VO_2_max) is measured using a spiroergometer (MedGraphics System Breese; MedGraphics, St. Paul, MN, USA). VO2max is expressed as VO2 per kg lean body mass (ml/min/kg).

### Gestational weight gain, birth outcome and offspring anthropometric data collection

As part of the standard care in Germany, gestational weight gain, birth outcome and offspring’s weight, length and head circumference are routinely and repeatedly documented in maternal and pediatric logs by health care providers. Data from these logs are recorded from all participants for study analyses.

### Questionnaires and food diary

During pregnancy, the Edinburgh Postnatal Depression Scale (EPDS) [38], which has also been validated for pregnant women [39], is used. The Beck Depression Inventory (BDI) II [40] is used as a psychometric screening test for depression in non-pregnant women. The German version of the Patient Health Questionnaire (PHQ-D) [41] is used to assess psychiatric disorders. Chronic stress is assessed with the Trier Inventory for the Assessment of Chronic Stress (TICS) [42]. Breastfeeding duration and intensity are assessed retrospectively with an in-house questionnaire (suppl. mat.). A 7-day food diary is collected from mothers and children at each visit and assessed with OptiDiet PLUS software Version 6.00.001 (GOE mbH, Linden, Germany). The maternal physical activity is assessed using the habitual physical activity (HPA) index [43] at each visit. In children physical activity is assessed with the MoMo-questionnaire [44] and pubertal status is self-assessed with a German pubertal questionnaire for children [45].

## Follow up of children

Offspring of PREG study participants are examined at age 2, 6, 10, 14 and 17 years (Table 1).

### Heart rate variability

In children, a 10-minute electrocardiogram is recorded with a BIOPAC MP36 (BIOPAC Systems Inc, Goleta, CA, USA) with a sampling rate of 1000 Hz. Children are positioned in a supine position and are instructed to lay still. In two-years old children, a caregiver is present and stillness is encouraged by reading a book. Time- and frequency-domain heart rate variability parameters are analyzed with KUBIOS HRV Software V2.2 [46].

### Developmental tests

In 2-year old children, the Bayley Scales of Infant Development Third Edition (BSID-III) [47] in the German Edition are carried out to assess the developmental state of the children. The BSID-III evaluates the development in infants aged 1 to 42 months in the domains cognition, language (receptive and active) and motor skills (fine and gross motor skills) on individual scales. The BSID-III also provides a social-emotional scale as well as an adaptive behavior scale, however, both were not available in German language at the beginning of the study and therefore were not included.

In order to achieve a high level of standardization, the developmental assessment of the children is done by a single researcher (JHa). A caregiver of the child is permanently present during measurement. Assessment with the BSID-III is always done in the morning and in the same specially prepared room. Disturbing factors due to environmental stimuli are largely excluded, and apart from the test material, the room is low in distractions to ensure the child’s attention is directed to the assessment.

### Intelligence assessment

The offspring intelligence is assessed with the Wechsler Intelligence Scale for Children – Fifth Edition (WISC-V) [48], which is one of the most implemented diagnostics to evaluate cognitive abilities in the age group of 6 to 16 years in children. The German Version of the WISC-V consists of a total of 15 subtests that can be used to generate standardized scores in 5 primary indices (verbal comprehension, visual spatial, fluid reasoning, working memory, processing speed), 5 secondary indices (quantitative reasoning, auditory working memory, nonverbal, general ability, cognitive proficiency) as well as a full-scale IQ score. The 15 subtests can be classified into two different categories, 10 primary and 5 secondary subtests. To build the full-scale IQ, the primary indices, and 3 of the 5 secondary indices, the 10 primary subtests are combined in different constellations. To ensure a standardized implementation, the WISC-V is conducted without the presence of parents by the same professional in a specially prepared room with a minimum of distractions.

### Physical activity

Daily physical activity is measured with a wrist accelerometer for children age 6 years and up for a time period of 14 days at each study visit (Fitbit Ace, Fitbit Inc., San Francisco, CA, US).

### Continuous glucose monitoring (CGM)

In children age 6 years and older at each visit 24h glucose profiles and postprandial glycemic excursions (AUC) will be measured by flash glucose monitoring (Freestyle Libre Pro iQ, Abbot Diabetes Care, Witney, UK). A single-use sensor is placed on the back of the upper arm of the child and stays in place for 14 days. Children and parents are unaware of the glucose levels, since the sensor is read out only after the recording period by study personnel.

### Patients and public involvement statement

Patients were not involved in the initial design of the PREG and PREG Offspring study. Annual meetings for study participants are conducted to connect patients with the responsible researchers. In addition, a newsletter with information on the latest research results is sent out once a year.

## Discussion and outlook

The prospective design of the PREG and PREG Offspring study with detailed phenotyping during and after pregnancy provides a unique platform to address crucial research questions to better understand the trajectories of subsequent maternal metabolism, fetal and offspring development and long-term health of mother and child. Several questions will be addressed. One key question is whether the standard GDM treatment starting after diagnosis in the third trimester of pregnancy is sufficient to prevent adverse effects on the offspring’s long-term health and development and thereby breaking the cycle of transgenerational diabetes.

The PREG study is also designed to examine the impact of maternal glucose excursions and other metabolic and anthropometric factors on the child’s brain development and some aspects of the proper formation of the autonomous nervous system. Tracking the development of cognition and intelligence as well as the development of the autonomous nervous system repeatedly throughout childhood and analyzing this in concert with fetal data obtained from fMEG/fMCG may provide unique insights on the effect of maternal metabolism on offspring’s neuronal development in humans [49].

Only a limited number of mother-child cohorts with focus on gestational diabetes exist worldwide, ranging from large register studies [50,51] over prospective studies [30,52–54] to case-control studies [50] and to follow up studies of randomized clinical trials for the prevention of GDM with lifestyle intervention early in pregnancy [55]. Though, the PREG study covers crucial aspects that are not comprehensively addressed in other larger trials so far. While register studies provide large datasets for epidemiological research, these studies lack the detailed characterization of subjects necessary for uncovering details of potential pathomechanisms of gestational diabetes. Prospective birth cohort studies usually recruit mother-child dyads only at birth and retrieve data on GDM diagnosis/OGTT from medical records, where often only potentially imprecise glucose measurements from baseline, 1h and 2h post glucose load are available. The PREG study is unique as it not only performs an OGTT with 5 strictly quality-controlled glucose measurements, but additionally insulin, C-peptide, proinsulin and free fatty acids are measured at each time point. Furthermore, biomaterial is properly stored for the measurement of upcoming novel parameters.

These detailed OGTT data will also be used to search for potential novel subtypes of GDM. Recently, novel clusters of prediabetic metabolism [56] and diabetes [57] have been defined in aiming for a better understanding of underlying disease pathomechanisms and prediction of disease progression and complications. These studies demonstrated that combining glucose levels with additional biomarkers can lead to a detection of clinically relevant metabolic subphenotypes [56]. The concept of subtyping of overt diabetes has been recently validated and their role for the early development of diabetes-related complications described [58]. Subclassification of GDM has been proposed before [59–61], but these studies relied only on glucose assessments, combined in part with insulin sensitivity. With such subclassification, a potential future T2DM risk may be captured more precisely but the clinical relevance of such approaches has yet to be proven in appropriate prospective studies. The comprehensive dataset collected from each pregnant woman in the PREG study will allow an investigation of possible subtypes of GDM with much higher discriminating power not only for the future diabetes risk of the mother but also for birth outcome and offspring’s development.

The unique strengths of the PREG study are the in-depth phenotyping of mothers during pregnancy and repeatedly after delivery, comprehensive laboratory measurements and state-of-the-art examination procedures in combination with biobanking. Data acquisition and sample handling are done according to standard operating procedures in all study sites, thus ensuring a high quality for each data point. Biosamples are collected during and after pregnancy which enables the measurement of potential new biomarkers or pathogenetic factors that might be identified in the future. The PREG Offspring study protocol provides a platform to comprehensively cover the period of childhood and adolescence. By design, the PREG study is not a population-based cohort but enriched for GDM cases.

In conclusion, the comprehensive phenotyping of maternal metabolism in this study will facilitate studying the effect of hyperglycemia and other maternal factors on subsequent diabetes risk of the mother and on body composition, metabolism and development of the offspring, with respect to standard GDM treatment. The findings could help adjust current treatment recommendations and by that contribute to fight the ever-increasing diabetes epidemic.

## Supporting information

suppl.mat.questionnaire.breastfeeding.and.infant.nutrition

## Data Availability

All requests for data and materials will be promptly reviewed by the Data Access Steering Committee to verify whether the request is subject to any intellectual property or confidentiality obligations. Individual-level data may be subject to confidentiality. Any data and materials that can be shared will be released via a Material Transfer Agreement. 

## Abbreviations

CGM: continuous glucose monitoring
fMEG: fetal magnetencephalography
fMCG: fetal magnetcardiography
GDM: gestational diabetes mellitus
MRI: magnetic resonance imaging
MRS: magnetic resonance spectroscopy
OGTT: oral glucose tolerance test
T2DM: type 2 diabetes mellitus

## Acknowledgments

We thank all of the volunteers for their participation in the study. We especially thank Vanessa Hartmann, Ines Wagener, Eva-Maria Stehle, Alexandra Eberle, Susanne Wegner, Dorothee Neuscheler and Henrike Peuker (all University of Tübingen) for their excellent technical assistance. We thank Sonja Hülskämper for the design of the graphical abstract.

## Author contributions

LF drafted the manuscript. LF, MH, JH, JHa, JHi, JM, KK, JPF, PJ, DL, RW performed examinations, LF, AP, SB, HP, HUH, MH, AS, MS, MHr, ALB, AF and MH contributed to concept of the study. The study was initiated by HUH. All authors contributed discussion of the manuscript and approved the final version before submission.

---

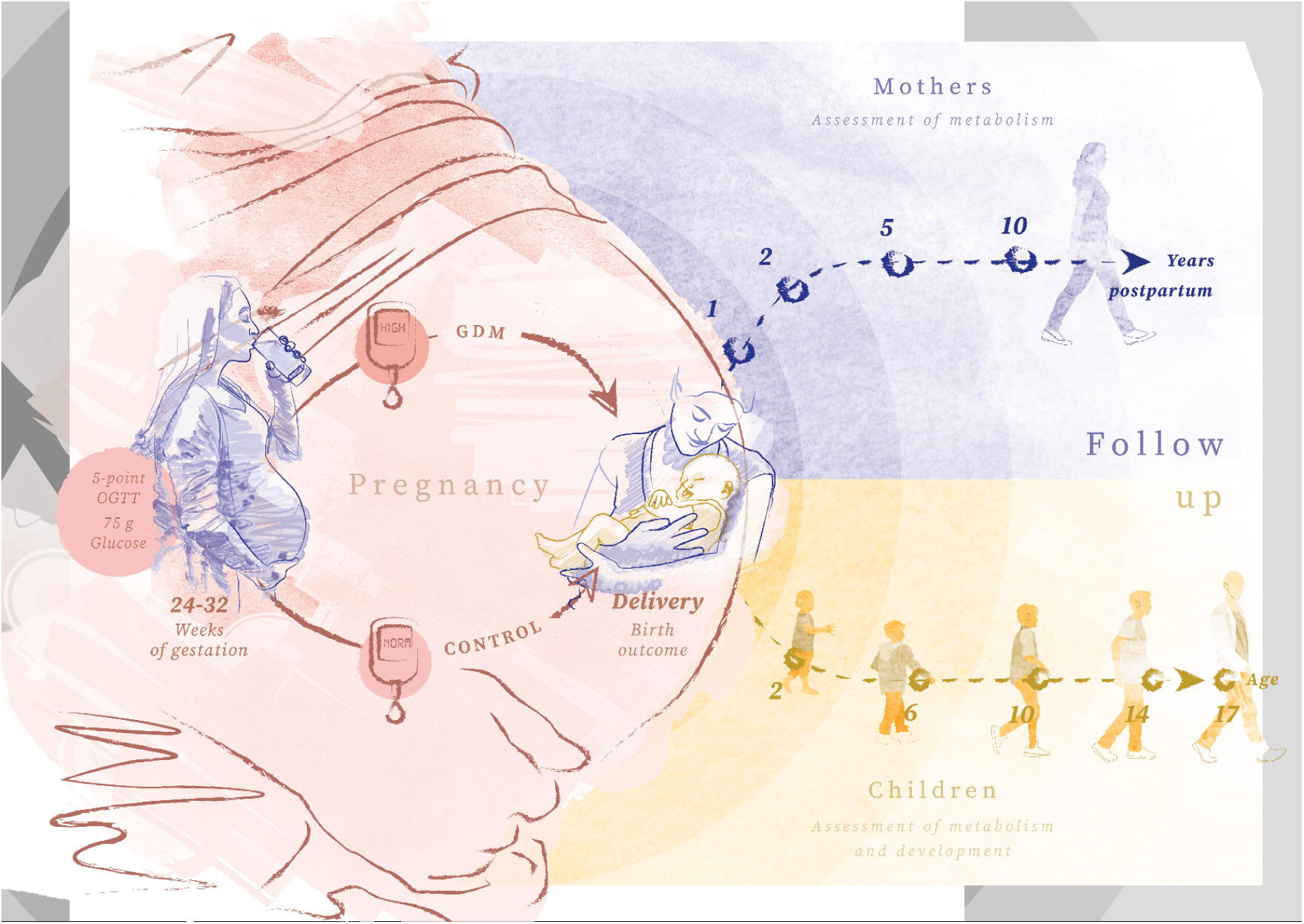

