## Supplementary material for "The German Gestational Diabetes Study (PREG): Rationale, Methodology and Design": Suppl.Tables

Table 1 Study sites PREG study

| Site | Examinations |
|---|---|
| **Tübingen**<br>Institute for Diabetes Research and Metabolic Diseases (IDM) at the Eberhard Karls University Tübingen | **Pregnancy:**<br>OGTT, liver sonography<br>**Delivery:**<br>Birth outcome, cord blood, placenta<br>**Follow up mothers:**<br>OGTT, spiroergometry, whole body MRI, MRS<br>**Follow up offspring:**<br>Developmental tests, intelligence test, heart rate variability, fasted blood, urine, whole body MRI, MRS, continuous glucose measurement |
| **Düsseldorf**<br>German Diabetes Center (DDZ), Leibniz Center for Diabetes Research at Heinrich Heine University Düsseldorf | **Pregnancy:**<br>OGTT<br>**Delivery:**<br>Birth outcome<br>**Follow up mothers:**<br>OGTT, spiroergometry, whole body MRI, MRS |
| **Leipzig**<br>University of Leipzig Medical Center, IFB Adiposity Diseases, University of Leipzig, | **Pregnancy:**<br>OGTT<br>**Delivery:**<br>Birth outcome<br>**Follow up mothers:**<br>OGTT, spiroergometry, whole body MRI, MRS |
| **Dresden**<br>Department of Medicine, University of Dresden | **Pregnancy:**<br>OGTT<br>**Delivery:**<br>Birth outcome<br>**Follow up mothers:**<br>OGTT, spiroergometry, abdominal MRI |

Table 2 LOINC codes for routinely measured analytes

| System | Analyte | Logical Observation Identifiers Names and Codes (LOINC) | Assessed in Mother (M) Child (C) |
|---|---|---|---|
| Serum/Plasma | Glucose | 2345-7 | M/C |
| | Insulin | 25447-4 | M/C |
| | C-peptide | 14633-2 | M/C |
| | Proinsulin | 27882-0 | M/C |
| | Non-esterified fatty acids | 1769-9 | M/C |
| | Leukocytes | 60026-2 | M/C |
| | Thrombocytes | 777-3 | M/C |
| | Erythrocytes | 789-8 | M/C |
| | Hematocrit | 4544-3 | M/C |
| | Hemoglobin | 718-7 | M/C |
| | Erythrocyte mean corpuscular hemoglobin (HBE) | 785-6 | M/C |
| | Erythrocyte mean corpuscular hemoglobin concentration (MCHC) | 786-4 | M/C |
| | Erythrocyte mean corpuscular volume (MCV) | 787-2 | M/C |
| | Albumin | 61152-5 | M |
| | Total protein | 2885-2 | M |
| | Triglycerides | 2571-8 | M/C |
| | Cholesterol | 2093-3 | M/C |
| | High density lipoprotein cholesterol (HDL-C) | 2085-9 | M/C |
| | Low density lipoprotein cholesterol (LDL-C) | 2089-1 | M/C |
| | Uric acid | 3084-1 | M |
| | Urea | 3091-6 | M |
| | Creatinine | 2160-0 | M |
| | Lipoprotein (a) | 10835-7 | M |
| | Apolipoprotein A-I | 1869-7 | M |
| | Apolipoprotein B | 1884-6 | M |
| | Homocysteine | 13965-9 | M |
| | Aspartate aminotransferase | 1920-8 | M |
| | Gamma glutamyl transferase | 2324-2 | M |
| | Alanine aminotransferase | 1742-6 | M |
| | HbA1c | 17856-6 | M/C |
| | Glutamate decarboxylase 65 Ab (GAD-AB) | 56540-8 | M |
| | Potassium | 2823-3 | M |
| | Sodium | 2951-2 | M |
| | Calcium | 2000-8 | M |
| | Chloride | 2075-0 | M |
| | Phosphate | 14879-1 | M |
| | Magnesium | 2601-3 | M |

| System | Analyte | Logical Observation Identifiers Names and Codes (LOINC) | Assessed in Mother (M) Child (C) |
|---|---|---|---|
| | Iron | 2498-4 | M |
| | Transferrin | 3034-6 | M |
| | C reactive protein (CRP) | 1988-5 | M/C |
| | Interleukin-6 (IL-6) | 26881-3 | M |
| | Thyrotropin (TSH) | 3016-3 | M |
| | Cortisol | 14675-3 | M |
| | β-human chorionic gonadotropin (β-HCG) | 19180-9 | M |
| | Prolactin | 2842-3 | M |
| | Estradiol | 14715-7 | M |
| | Progesterone | 14890-8 | M |
| | Testosterone | 14913-8 | M |
| | Sex hormone binding protein (SHBG) | 13967-5 | M |
| Urine | Albumin | 1754-1 | M/C |
| | Creatinine | 2161-8 | M/C |
| | Urea | 3092-4 | M/C |
| | Total protein | 2888-6 | M/C |
| | Leukocytes | 33052-2 | M/C |
| | Nitrit | 50558-6 | M/C |
| | pH | 50560-2 | M/C |

Table 3 Grading of hepatic steatosis

| Grade | Echogenicity | Sound attenuation | Vessels |
|---|---|---|---|
| I | + | - | Normal |
| II | ++ | (+) | Normal |
| III | +++ | + | Rarified |
