## Supplementary material for "The German Gestational Diabetes Study (PREG): Rationale, Methodology and Design": suppl.mat.questionnaire.breastfeeding.and.infant.nutrition

*Please ONLY answer the questions for the infant included with you in the study.*

My Child was born on: ____. ____. ________ (dd/mm/yy)

---

**1. How did you feed your child before you implemented the supplementary diet?**

Before I implemented the supplementary diet I

- ☐ breastfed exclusively (only breast milk) → *continue with question 3*
- ☐ breastfed partially (breast milk + infant formula)
- ☐ did not breastfeed.................................................. → *continue with question 4*
- ☐ breastfed for a very short period (< 1 week) ............... → *continue with question 5*
- ☐ am still breastfeeding

**2. If you used to breastfeed partially after delivery, which of the following statements describes the ratio between breast milk and baby milk best?**

- ☐ I mainly breastfed (approx. 75% breast milk and 25% infant formula)
- ☐ I equally fed breast milk and infant formula (approx. 50% breast milk and 50% infant formula)
- ☐ I mainly fed infant formula (approx. 25% breast milk and 75% infant formula)

3. **For how long have you been breastfeeding your infant in total?** (Pleas also consider the months you have been breastfeeding partially, e.g. during the start of supplementary diet)

   - I have been breastfeeding my infant for ☐☐ months in total
   - During this time, I have been breastfeeding

     ☐☐ months exclusively (only breast milk)

     ☐☐ months partially (breast milk + infant formula / supplementary food))

**→ If you used to breastfeed exclusively or partially please continue with question 6**

---

**Please answer the next question only if you did <u>not breastfeed</u> your infant at all**

4. **What was the reason why you chose not to breastfeed your infant?** (multiple answers possible)

   - ☐ I did not want to
   - ☐ I was sick and / or had to take medication
   - ☐ My baby had health issues (e.g. neonatal jaundice)
   - ☐ I had problems with breastfeeding my infant (e.g. unfavorable nipples)
   - ☐ I had a lack of knowledge about how to breastfeed correctly
   - ☐ I was concerned about foreign substances in the breast milk
   - ☐ I wanted the possibility to smoke / drink again
   - ☐ I thought bottle feeding is more comfortable / handier
   - ☐ I did not have enough time / I had too much stress
   - ☐ I had to / wanted to go to work again
   - ☐ My infant had problems with suckling / refuse to nurse/breastfeed
   - ☐ I did not have enough breast milk
   - ☐ I wanted to have my body back to myself
   - ☐ I felt uncomfortable / embarrassed while breastfeeding
   - ☐ Other reasons:_____________________________________________

**Please answer the next question only if you breastfed for less than one week**

5. **What was your reason to quit breastfeeding?** (multiple answers possible)

- ☐ I got sick and / or had to take medication
- ☐ My child got health issues (e.g. neonatal jaundice)
- ☐ I had problems with breastfeeding my infant (e.g. mastitis)
- ☐ I was concerned about foreign substances in the breast milk
- ☐ I wanted the possibility to smoke / drink again
- ☐ I thought bottle feeding is more comfortable / handier
- ☐ I did not have enough time / I had too much stress
- ☐ I had to / wanted to go to work again
- ☐ My infant had problems with suckling / refuse to nurse
- ☐ My breast milk supply was not sufficient / my infant did not gain enough weight
- ☐ I wanted to have my body back to myself
- ☐ I felt uncomfortable / embarrassed while breastfeeding
- ☐ I did not feel advised sufficiently / I was to insecure how to breastfeed correctly
- ☐ I thought it was the right time to quit breastfeeding/ For me it was the right time to end breastfeeding
- ☐ Other reasons:_______________________________________________

---

6. **Since I started the supplementary diet - besides breast milk/ infant formula and water - my child also got the following beverages in the first two years of life:** (multiple answers possible)

- ☐ unsweetened tea
- ☐ sweetened tea
- ☐ fruit juice
- ☐ nothing additional to breast milk/baby milk and water
- ☐ others:_______________________________________________

**7. When did you implement supplementary diet?**

- I started feeding supplementary diet at the ☐☐ months
- I used to breastfeed additionally during the start of supplementary diet:

☐ Yes ☐ No

**8. Which of the following statements describes your ratio between homemade and store-bought baby food best?**

☐ I only fed homemade baby food

☐ I fed homemade baby food mainly (approx. 75% homemade baby food)

☐ I equally fed breast milk and infant formula (approx. 50% homemade and 50% store-bought baby food)

☐ I mainly fed store-bought baby food (ca. 75% baby food)

☐ I only fed store-bought baby food

**9. Did your child get additional „snacks“ (e.g. millet rings, squeeze pouches) in the first two years of life?**

☐Yes ☐No

If yes, what kind of snacks did they get? ________________________________________

If yes, how often did they get such snacks?

☐ daily

☐ 5-6 times per week

☐ 3-4 times per week

☐ 1-2 times per week

☐ rarely / only in special situations

**10. Did you actively change your lifestyle (your nutrition and / or activity) during the first two years after delivery in order to get fitter or reach your desired weight?**

☐ Yes ☐ No

If yes: what did you change?

☐ I changed my general nutrition (e.g. started vegetarian / vegan diet)

☐ I started a diet (especially for weight reduction, e.g. low carb, weight watchers)

☐ I was more active (e.g. went for a walk more often)

☐ I was more active <u>and</u> changed my nutrition

☐ I started a new sport discipline

☐ others: ______________________________________________

**Thank you for your participation!**
